# The effects of walking in nature on negative and positive affect in adult psychiatric outpatients with major depressive disorder: A randomized-controlled study

**DOI:** 10.1101/2021.11.25.21266872

**Authors:** Kia Watkins-Martin, Despina Bolanis, Stéphane Richard-Devantoy, Marie-Hélène Pennestri, Catherine Malboeuf-Hurtubise, Frederick Philippe, Julie Guindon, Jean-Philippe Gouin, Isabelle Ouellet-Morin, Marie-Claude Geoffroy

**Affiliations:** Department of Educational and Counselling Psychology, McGill University, Montreal, QC, Canada; Department of Psychiatry, McGill University, Montreal, QC, Canada; Department of Psychiatry, Saint-Jérôme Hospital, Saint-Jérôme, QC, Canada; Hôpital en santé mentale Rivière-des-Prairies (CIUSSS-NIM), Montreal, QC, Canada; Department of Psychology, Bishop’s University, Sherbrooke, QC, Canada; Department of Psychology, Université du Québec à Montréal, Montreal, QC, Canada; Department of Psychology, Concordia University, Montreal, QC, Canada; School of Criminology, University of Montreal, Montreal, QC, Canada; Research Center of the Montreal Mental Health University Institute, Montreal, QC, Canada

**Keywords:** major depressive disorder (MDD), depression, nature, greenspace, affect, randomized-control trial, physical activity

## Abstract

**Background:** While walking in nature has been shown to improve affect in adults from the community to a greater extent than walking in urban settings, it is unknown whether such findings can be generalized to individuals suffering from depression. Using a parallel group design, this randomized controlled trial examined the effects of a single walk in nature versus urban settings on negative and positive affect in adult psychiatric outpatients diagnosed with major depressive disorder (MDD).

**Method:** Participants recruited from a psychiatric outpatient clinic for adults with MDD were randomly assigned to a nature or urban walk condition. Thirty-seven adults (mean age=49 years) completed a single 60-minute walk. Negative and positive affect were assessed using The Positive and Negative Affect Schedule or PANAS at 6 time points: before the walk, halfway during the walk, immediately post-walk, at home before bedtime, 24 hours post-walk, and 48 hours post-walk.

**Results:** Controlling for baseline levels of affect before the walk, individuals who walked in nature experienced overall lower levels of negative affect, *F*(1, 35.039)=4.239, *p*=.047, compared to those who walked in urban settings. Positive affect did not differ across walk conditions.

**Limitations:** The generalizability of results are limited by the small sample size and the presence of more female than male participants.

**Conclusions:** Walking in nature might be a useful strategy to improve the affect of adults with MDD. Future research should investigate different ways to integrate the beneficial effects of nature exposure into existing treatment plans for psychiatric outpatients with MDD.

## Introduction

Depression is a leading cause of disability and one of the most common mental disorders, affecting over 300 million people worldwide (Friedrich, 2017; Vos et al., 2015; World Health Organization, 2017). While psychotherapy, pharmacotherapy, or a combination of both are effective in reducing depressive symptoms (Cuijpers, 2014; Kamenov et al., 2017), there remains a need for free, complementary strategies that can be easily implemented by individuals without professional assistance (Ravindran et al., 2016).

In recent years, there has been a growing interest in the benefits of spending time in nature for mental health (Bowler et al., 2010; Bragg and Atkins, 2016; Bratman et al., 2021; Britton et al., 2020; Frumkin et al., 2017; Hartig and Kahn, 2016; Keniger et al., 2013; Kondo et al., 2018; White et al., 2017), including the alleviation of depression symptoms (Bratman et al., 2019; Sarkar et al., 2018; White et al., 2021). One practical way to spend time in nature is to walk, a free and highly accessible form of physical activity that most people find enjoyable. Within the general population, a growing number of studies have reported that walking in nature confers greater benefits for affect than walking in urban settings (Berman et al., 2008; Bratman et al., 2015a; Hartig et al., 2003; Janeczko et al., 2020; Koselka et al., 2019). To illustrate, Bratman and colleagues (2015a) found that healthy young adults (mean age=22.9 years) who walked in nature (*n*=30) for 60 minutes reported a greater decrease in negative affect and increase in positive affect from before to immediately after the walk compared to those who walked in an urban environment (*n*=30) for the same duration. To our knowledge, only one prior study has examined whether a single walk in nature can improve the affect (i.e., decrease negative affect and increase positive affect) of adults diagnosed with depression. Using a within-subjects randomized crossover design with 19 adults diagnosed with depression, a study by Berman and colleagues (2012) found that positive affect increased to a greater extent after a 50-minute nature walk vs. an urban walk alongside a busy public road, whereas negative affect improved (i.e. decreased) for all participants, regardless of walk condition.

The present study aims to evaluate the effects of a single walk in nature (i.e., in a large, biodiverse park) versus urban settings (i.e., alongside a busy public road) on levels of negative and positive affect in adult psychiatric outpatients with MDD, using a rigorous randomized controlled protocol. Clarifying whether such findings can be replicated in a population of psychiatric outpatients with MDD could inform the development of complementary strategies to alleviate depressive symptoms. While virtually all prior studies comparing the effects of a single nature vs. urban walk (Berman et al., 2012, 2008; Bratman et al., 2015a; Janeczko et al., 2020) only measured affect immediately before and after the walk, our study will further extend prior knowledge by using repeated assessments of affect, including measures at mid-point during the walk; immediately after the walk; and at several points post-walk (the same day before going to bed; the next morning; and two days later).

## Method

### Ethics Statement

The study was approved by the ethics board of the Douglas Mental Health University Institute. Participants provided written informed consent and were compensated up to $75 for their participation. This randomized controlled trial (RCT) using parallel group design is registered at ClinicalTrials.gov (NCT03996785). Changes made to the study protocol due to the COVID-19 pandemic are detailed in the supplemental material.

### Participants

A total of 47 participants were recruited in 2019 and 2021 from a tertiary psychiatric outpatient clinic for adults with refractory major depressive disorder, most of the time accompanied by suicidal ideation, at the Douglas Mental Health University Institute (DMHUI) in the province of Quebec, Canada. Patients were eligible to participate in this study if they were between the ages of 18 to 65 years old; had a primary diagnosis of MDD (as per their medical chart according to the Structured Clinical Interview for Axis I DSM-IV, SCID I; First, 1997); were able to walk for one hour; had the ability to complete online follow-up questionnaires; and were fluent in English or French. Participants were excluded if they had imminent suicidal risk (i.e., desire and intent, as well as the capacity of carrying out intent within 48 hours); a lifetime history of psychotic disorders; or a heart or serious medical condition (e.g., major surgery) that might impede their ability to walk for an hour. At the time of the trial, all participants were receiving care from the psychiatric hospital’s medical team, which includes psychiatrists, nurses, psychologists, and social workers.

Of the 47 recruited participants, 10 dropped out prior to completing their scheduled walk and 37 completed the walk (see **Figure 1**). Participants who completed the walk and were included in the analyses (*n*=37) were not different from those who dropped out prior to their scheduled walk (*n*=10) in terms of sex (*p*=.889), age (*p*=.729), and severity of depressive symptoms at baseline (*p*=.856) assessed using the Hamilton Depression Rating Scale (Hamilton, 1967). Among the 37 participants who completed their walk (including the first three assessments: Pre-Walk, During the Walk, and Immediately Post-Walk), 32 participants completed the assessments at Time 4 (Before Bedtime) and 5 (24 Hours Post-Walk) and 29 participants completed the assessments at all 6 time points (including Time 6, 48 Hours Post-Walk). Of the 37 participants who completed the walk, 20 were randomized to the nature walk condition and 17 to the urban walk condition.

**Figure 1.**
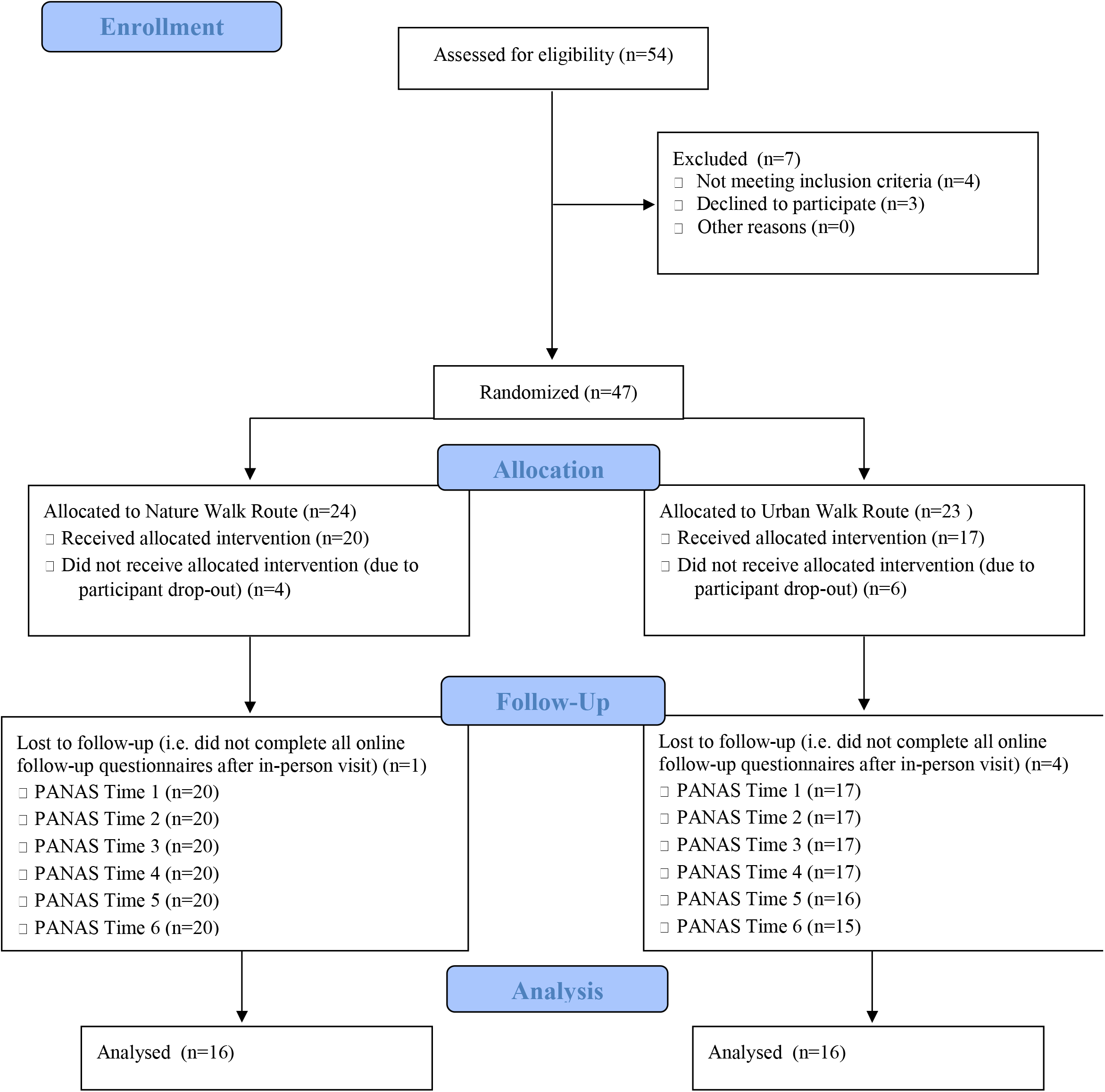
CONSORT diagram of participants’ progress through the study.

### Randomization

Participants were randomized to the condition groups (nature walk vs. urban walk) using a computerized randomization procedure (www.keamk.com) with a 1:1 allocation ratio. This study is a single-blind RCT; as such, only the participants were blind to the randomization conditions. In the process of obtaining informed consent, participants were told that the goal of the study was to examine the impact of different outdoor walk routes on depression. The research assistants conducting the assessments and guiding the walks were not blind to participants’ assigned walk condition.

### Walk conditions

Walks took place from May 20^th^ to October 23^rd^ in 2019 and from June 21^st^ to August 20^th^ in 2021 in the morning, between 8am and 12pm (the study was interrupted in 2020 due to the COVID-19 pandemic). Participants went on one scheduled walk, individually or in groups of two, while accompanied by two research assistants (psychology undergraduate/graduate students). They walked for approximately 60 minutes at a pace of 3-5km/hour and were instructed to refrain from engaging in conversation to minimize social stimulation and maximize their focus on the surrounding environment. The settings and duration of the nature and urban walk conditions described below were similar to those used in prior studies (Berman et al., 2012, 2008; Bratman et al., 2015a) to increase inter-study comparability. In the case of heavy rain, walks were rescheduled to the participants’ next availability. Information on the weather (sunny / partly sunny, cloudy / partly cloudy) was obtained via Environment Canada (see **Supplemental Table 1**). Both the nature and urban walk conditions were similar in terms of weather conditions (recorded from Environment Canada) and perceived levels of physical exertion, as reported by the BORG CR-10 scales (Borg, 1998) completed by participants immediately after each walk (participants were asked to answer the question “How was the walk?” by circling a number on a scale ranging from 0-*No exertion at all* to 10-*Maximal exertion*; see **Supplemental Table 1**).

The nature walk took place in a 97-hectare biodiverse urban park near the psychiatric hospital. The total distance of the nature walk was 4.41km with a cumulative elevation of 13 meters and lasted approximately 60 minutes. The distance between the psychiatric clinic building entrance to the park entry was 0.92km, 0.77km of which was spent crossing the clinic parking lot. As such, participants walked for about 14 minutes before reaching the park entrance. The walk included a stretch of 3.5 km within the park far from urban sounds (e.g., automobile noise) and sights (e.g., buildings and parked automobiles). The park consists of a forest of with over 20,000 trees as well as a pond, and no automobiles are permitted within the vicinity of the park.

The urban walk took place on the sidewalks of the busiest street near the psychiatric hospital with 3-4 lanes of automobile traffic. Participants walked west on this street and completed a short loop at the midpoint of the walk within the residential streets and then turned back around, in order to match the distance of the nature walk (see **Figure 2**). The total distance of the urban walk was 4.46 km with a cumulative elevation of 8 meters. Participants walking on this route were exposed to a significant amount of noise generated by automobiles.

**Figure 2.**
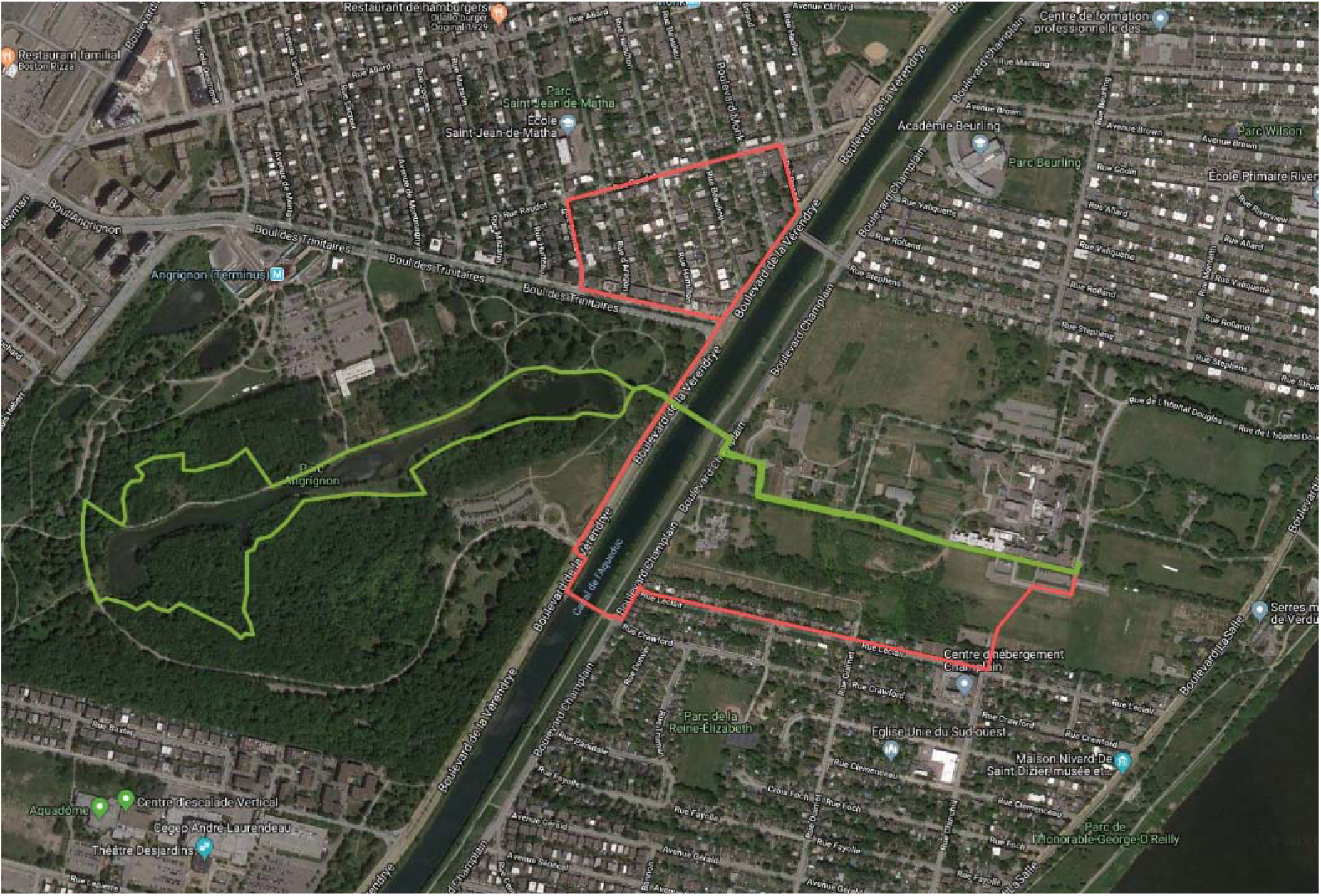
GoogleMaps overview of the nature walk condition and the urban walk condition. *Note*: The nature condition walk route is highlighted in green. The urban condition walk route is highlighted in red.

### Assessments

Following randomization, the assessments first took place at the hospital in the hour preceding the scheduled walk (Time 1, “Pre-Walk”); during the walk at midpoint (Time 2, “During the Walk”); immediately after the walk at the hospital (Time 3, “Immediately Post-Walk”); later in the evening at home (Time 4, “Before Bedtime”); the morning after the walk (Time 5, “24 Hours Post-Walk”); and 48 hours after the walk (Time 6, “48 Hours Post-Walk”).

### Primary and secondary outcomes

The study’s primary and secondary outcomes were negative and positive affect, respectively, as measured by the Positive and Negative Affect Schedule (PANAS; Watson et al., 1988). The PANAS is a widely-used 20-item self-rated questionnaire measuring positive (e.g., “excited”) and negative (e.g., “upset”) affect (positive affect scale, α: 0.88-0.93; negative affect scale, α: 0.89-0.93). Participants indicated to what extent they feel each affect “right now—that is, at the present moment” with response choices ranging from 1 (*very slightly or not at all)* to 5 (*extremely)*. The PANAS was administered via iPad at all 6 time points of the study: Pre-Walk (Time 1), During the Walk (Time 2), Immediately Post-Walk (Time 3), Before Bedtime (Time 4), 24 Hours Post-Walk (Time 5), and 48 Hours Post-Walk (Time 6).

### Statistical Analyses

First, we compared participants assigned to the nature vs. urban walk condition on baseline sociodemographic and clinical characteristics using 2-tailed T-tests for continuous measures and chi-square tests for categorical measures. Second, we examined differences in both negative and positive affect between walk conditions using 2-tailed T-tests. Third, we evaluated the effects of walk condition on negative and positive affect using two repeated measure mixed model analyses with Group (i.e. walk condition; nature vs. urban) as the between-subject factor, Time as the within-subject factor. Mixed models are ideal for analyzing repeated measures data and can accommodate for missing data via the Full Information Maximum Likelihood (FIML) method while using all available observed measurements (Detry and Ma, 2016; Enders, 2001). All analyses were conducted using SPSS, version 28, and the threshold of significance was set at *p* >.05.

## Results

As shown in **Table 1**, participants in both groups were similar in terms of baseline characteristics, including the severity of depressive symptoms assessed by the HAM-D (Hamilton, 1967). However, differences in negative (but not positive) affect between groups were detected at all time points, including at Time 1 (Pre-Walk; *p*=.022), with participants assigned to the nature walk condition showing lower levels of negative affect at baseline than those assigned to the urban walk (see **Table 2**). Thus, Pre-Walk affect (both positive and negative) was controlled for in all statistical models.

**Table 1.**
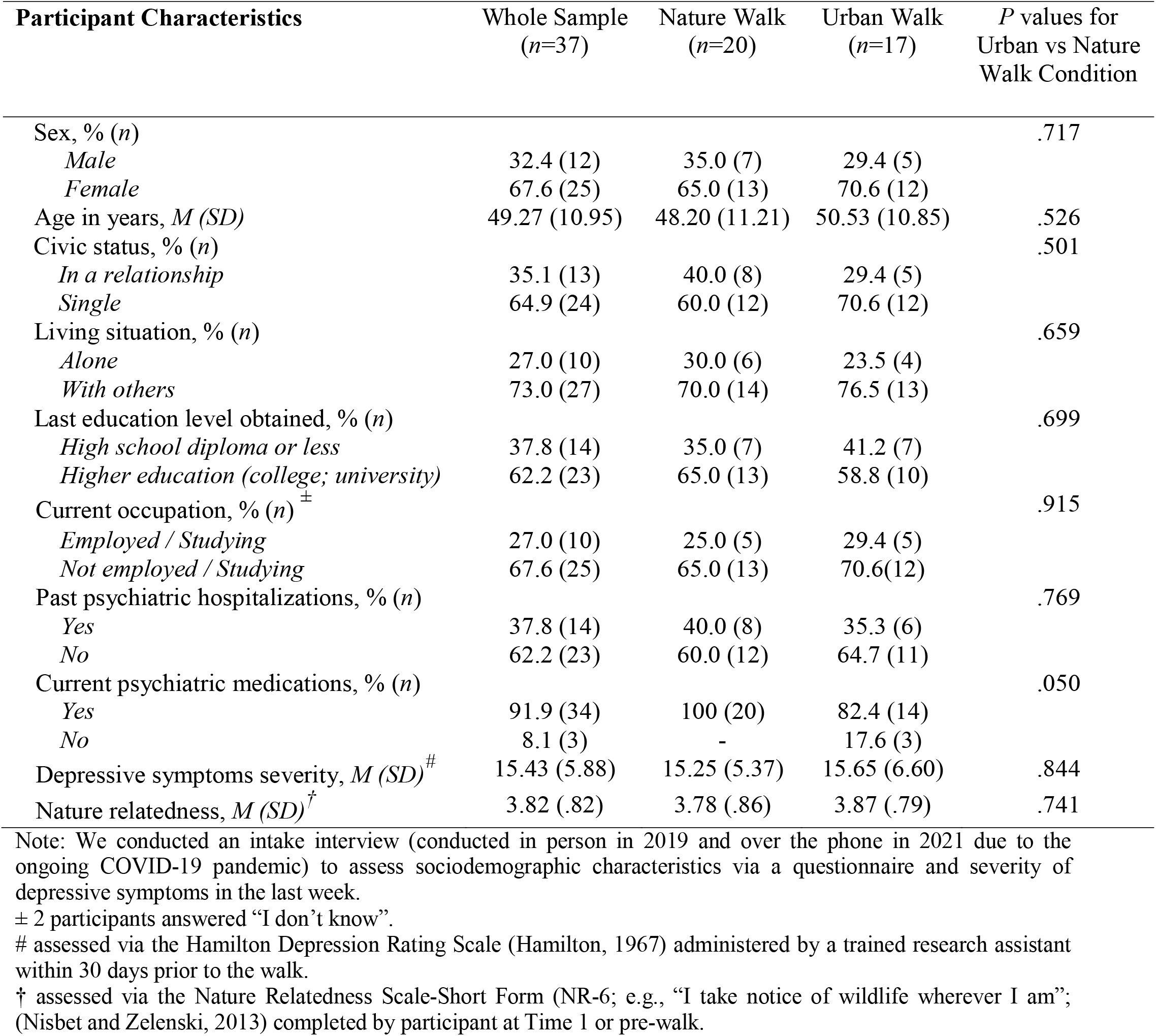
Baseline sociodemographic, and clinical characteristics of participants by walk condition.

| **Participant Characteristics** | **Whole Sample**<br>**(*n*=37)** | **Nature Walk**<br>**(*n*=20)** | **Urban Walk**<br>**(*n*=17)** | ***P* values for**<br>**Urban vs Nature**<br>**Walk Condition** |
| --- | --- | --- | --- | --- |
| Sex, % (*n*) |  |  |  | .717 |
| *Male* | 32.4 (12) | 35.0 (7) | 29.4 (5) |  |
| *Female* | 67.6 (25) | 65.0 (13) | 70.6 (12) |  |
| Age in years, *M* (*SD*) | 49.27 (10.95) | 48.20 (11.21) | 50.53 (10.85) | .526 |
| Civic status, % (*n*) |  |  |  | .501 |
| *In a relationship* | 35.1 (13) | 40.0 (8) | 29.4 (5) |  |
| *Single* | 64.9 (24) | 60.0 (12) | 70.6 (12) |  |
| Living situation, % (*n*) |  |  |  | .659 |
| *Alone* | 27.0 (10) | 30.0 (6) | 23.5 (4) |  |
| *With others* | 73.0 (27) | 70.0 (14) | 76.5 (13) |  |
| Last education level obtained, % (*n*) |  |  |  | .699 |
| *High school diploma or less* | 37.8 (14) | 35.0 (7) | 41.2 (7) |  |
| *Higher education (college; university)* | 62.2 (23) | 65.0 (13) | 58.8 (10) |  |
| Current occupation, % (*n*)<sup>±</sup> |  |  |  | .915 |
| *Employed / Studying* | 27.0 (10) | 25.0 (5) | 29.4 (5) |  |
| *Not employed / Studying* | 67.6 (25) | 65.0 (13) | 70.6 (12) |  |
| Past psychiatric hospitalizations, % (*n*) |  |  |  | .769 |
| *Yes* | 37.8 (14) | 40.0 (8) | 35.3 (6) |  |
| *No* | 62.2 (23) | 60.0 (12) | 64.7 (11) |  |
| Current psychiatric medications, % (*n*) |  |  |  | .050 |
| *Yes* | 91.9 (34) | 100 (20) | 82.4 (14) |  |
| *No* | 8.1 (3) | - | 17.6 (3) |  |
| Depressive symptoms severity, *M* (*SD*)<sup>#</sup> | 15.43 (5.88) | 15.25 (5.37) | 15.65 (6.60) | .844 |
| Nature relatedness, *M* (*SD*)<sup>†</sup> | 3.82 (.82) | 3.78 (.86) | 3.87 (.79) | .741 |

Note: We conducted an intake interview (conducted in person in 2019 and over the phone in 2021 due to the ongoing COVID-19 pandemic) to assess sociodemographic characteristics via a questionnaire and severity of depressive symptoms in the last week.
± 2 participants answered “I don’t know”.

**Table 2.** Means and standard deviations for positive and negative affect scores in the urban group and nature group (*n*=32) at Pre-Walk (Time 1), During the Walk (Time 2), Immediately Post-Walk (Time 3), Before Bedtime (Time 4), 24 Hours Post-Walk (Time 5) and, 48 Hours Post-Walk (Time 6).

| Time | Negative Affect |  |  | Positive Affect |  |  |
| --- | --- | --- | --- | --- | --- | --- |
|  | Nature Group<br>*M* (SD) | Urban Group<br>*M* (SD) | *P* Values | Nature Group<br>*M* (SD) | Urban Group<br>*M* (SD) | *P* Values |
| Pre-Walk <sup>1</sup> | 18.80 (6.80) | 25.06 (9.07) | .022 | 22.95 (7.58) | 23.06 (7.60) | .966 |
| During the Walk <sup>2</sup> | 13.45 (3.61) | 17.77 (7.63) | .031 | 25.80 (8.65) | 28.59 (6.97) | .294 |
| Immediately Post-Walk <sup>3</sup> | 12.00 (2.69) | 17.11 (6.96) | .005 | 24.45 (10.24) | 27.18 (7.28) | .565 |
| Before Bedtime <sup>4</sup> | 13.78 (5.22) | 21.00 (8.65) | .005 | 21.72 (9.34) | 22.41 (7.22) | .809 |
| 24 Hours Post-Walk <sup>5</sup> | 15.00 (5.37) | 23.25 (7.66) | <.001 | 20.44 (9.07) | 22.06 (9.26) | .611 |
| 48 Hours Post-Walk <sup>6</sup> | 16.28 (6.94) | 23.33 (9.12) | .017 | 20.67 (7.82) | 21.13 (6.88) | .858 |

Note. Based on maximum $n$ available for each time point.
<sup>1</sup> Nature Group, $n=20$ ; Urban Group, $n=17$
<sup>2</sup> Nature Group, $n=20$ ; Urban Group, $n=17$
<sup>3</sup> Nature Group, $n=20$ ; Urban Group, $n=17$
<sup>4</sup> Nature Group, $n=18$ ; Urban Group, $n=17$
<sup>5</sup> Nature Group, $n=18$ ; Urban Group, $n=16$
<sup>6</sup> Nature Group, $n=18$ ; Urban Group, $n=15$

We performed two repeated measure mixed model analyses to test the effects of a single walk in nature vs. urban settings on negative and positive affect while controlling for baseline affect at Time 1 (Pre-Walk). For negative affect, results indicated that there was no significant interaction between Group and Time on negative affect, *F*(4, 128.729) = 1.223, *p*=.304, and no main effect of Time, *F*(4, 128.736) = 0.471, *p*=.757. However, there was a significant main effect of group, *F*(1, 35.039) = 4.239, *p*=.047. As illustrated in **Table 3** and **Figure 3**, after adjusting for Pre-Walk (Time 1) levels of negative affect, participants who walked in nature reported overall lower levels of negative affect than those who walked in urban settings. For positive affect, results indicated no statistically significant Group x Time interaction, *F*(4, 127.131)=0.752, *p*=.559, and no significant main effect of Group, *F*(1, 33.662)=0.393, *p*=.535. However, there was a significant main effect of time on positive affect, *F*(4, 127.556)=6.059, *p*<.001 (see **Table 3** and **Figure 4**).

**Table 3.** Repeated measures mixed model summary table examining the effects of walking in nature vs. urban settings on negative and positive affect over time, while adjusting for Pre-Walk (Time 1) levels of negative and positive affect; *n*=37.

| Factor | Negative Affect |  |  | Positive Affect |  |  |
| --- | --- | --- | --- | --- | --- | --- |
|  | *df* | *F* | *p* | *df* | *F* | *p* |
| Time | 4, 128.736 | .471 | .757 | 4, 127.556 | 6.059 | **<.001** |
| Group | 1, 35.039 | 4.239 | **.047** | 1, 33.662 | .393 | .535 |
| Time x Group | 4, 128.729 | 1.223 | .304 | 4, 127.131 | .752 | .559 |

*Note.*
Covariate: Time 1 (Pre-Walk).
Time: Time 2 (During the Walk); Time 3 (Immediately Post-Walk); Time 4 (Before Bedtime); Time 5 (24 Hours Post-Walk); and Time 6 (48 Hours Post-Walk).
Group: Nature walking condition; Urban walking condition

**Figure 3.**
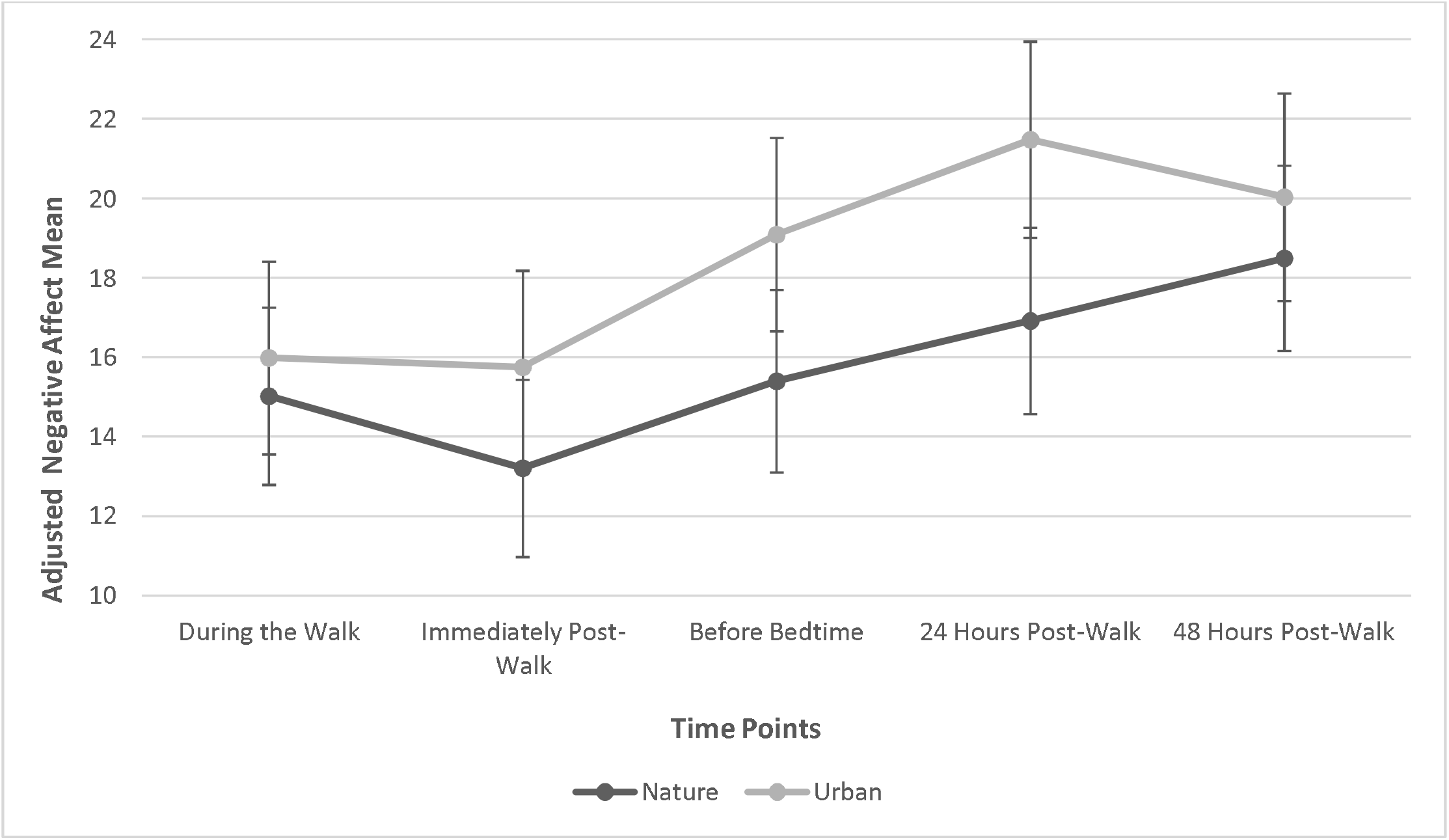
Adjusted mean negative affect levels During the Walk (Time 2), Immediately Post-Walk (Time 3), Before Bedtime (Time 4), 24 Hours Post-Walk (Time 5), and 48 Hours Post-Walk (Time 6), while controlling for Pre-Walk (Time 1); *n*=37.

**Figure 4.**
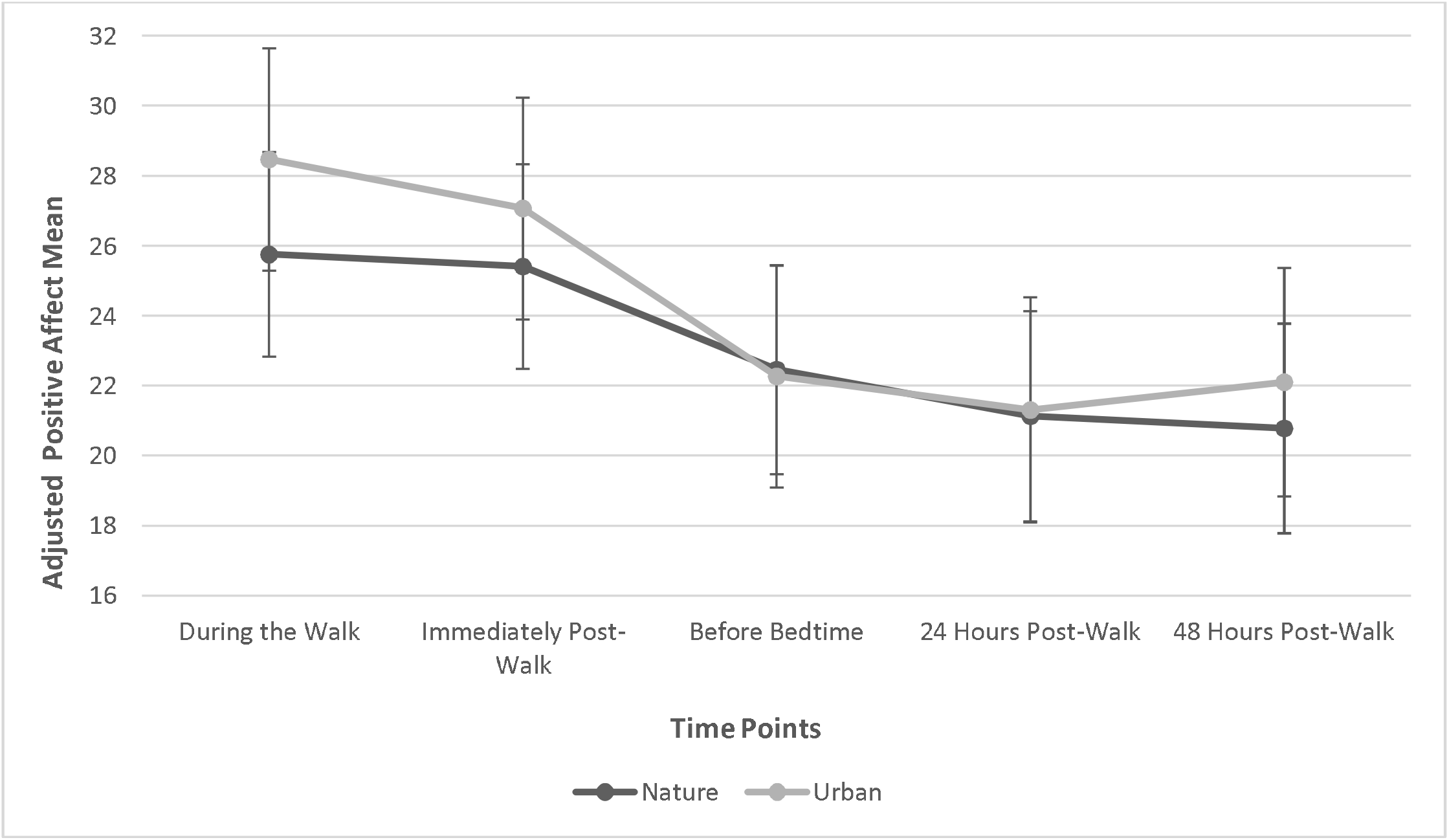
Adjusted mean positive affect levels During the Walk (Time 2), Immediately Post-Walk (Time 3), Before Bedtime (Time 4), 24 Hours Post-Walk (Time 5), and 48 Hours Post-Walk (Time 6), while controlling for Pre-Walk (Time 1); *n*=37.

## Discussion

Using a robust RCT design, this study suggests that walking in nature (vs. in urban settings) can lower negative affect in adults with MDD recruited from a psychiatric outpatient clinic for refractory depression. Our results showed that after controlling for baseline differences in affect, participants who walked in nature experienced less negative affect overall (and no difference in positive affect) compared to those who walked in an urban setting.

Overall, our findings indicating lower levels of negative affect in participants who walked in nature relative to those who walked along an urban route are in line with results from other experimental studies conducted with non-depressed populations (Berman et al., 2008; Bratman et al., 2015a; Janeczko et al., 2020; Koselka et al., 2019). To our knowledge, the 2012 study by Berman and colleagues is the only other study that has examined the effect of a single walk in nature vs. urban settings on both negative and positive affect of young adults diagnosed with MDD (*n*=19; 12 females; mean age=26 years; Berman et al., 2012). Unlike our findings indicating lower levels of overall negative affect (with no difference in positive affect) among participants who walked in nature, Berman and colleagues’ results showed a greater improvement in positive affect among participants who walked in nature (vs. in an urban setting), with no group difference in terms of negative affect. It is worth noting that participants in Berman and colleagues’ study were primed to ruminate about a negative autobiographical experience just prior to the walk, which may have interfered with anticipated benefit of nature exposure on participants’ negative affect. Further, although Berman’s participants were diagnosed with MDD, they were recruited from the community, not a psychiatric hospital. It is plausible that our participants, all psychiatric outpatients with a formal diagnosis of MDD, showed changes in affect that are typical of individuals with more severe depression. Indeed, prior studies have shown that individuals suffering from depression report lower levels of positive emotion compared to non-depressed people during exposure to positive stimuli (Berenbaum and Oltmanns, 1992; Bylsma et al., 2008; Horner et al., 2014; Pizzagalli et al., 2008; Rottenberg, 2005; Rottenberg et al., 2002; Sloan et al., 1997; Vanderlind and Joormann, 2019) and the presence of greater depressive symptoms is correlated with higher levels of positive emotion dampening (i.e., active or passive attempts to downregulate positive emotion after it has been elicited; Eisner et al., 2009; Feldman et al., 2008; Nelis et al., 2015).

### Strengths and Limitations

To the best of our knowledge, our study is the first to examine the effects of a single walk in nature (vs. urban settings) on the affect of an adult psychiatric outpatient population with MDD. Other strengths include its’ single-blind RCT study design; the use of a standardized measure of positive and negative affect; the prospective data collection at six points in time; and the use of an urban park (i.e., a large, biodiverse park found in an urban setting) to assess the impact of nature exposure while walking, as urban parks are likely to be more accessible to city residents than countryside settings. We nevertheless acknowledge the following limitations. Our preliminary analyses revealed baseline differences in levels of negative affect between participants in the nature vs. urban walk conditions (there was no observed baseline difference in positive affect). In this single-blind RCT, participants were blind to their assigned walk condition – indeed, while they were told in the process of obtaining informed consent that the goal of the study was to examine the impact of different outdoor walk routes on depression, participants were not made aware of the explicit aim to compare the effects of walking in nature vs. urban settings. While the research assistants administering assessments and guiding the walks were not blind to participants’ assigned walk condition, the baseline assessment of affect was administered via iPad to participants within the first 15 minutes of their arrival to the testing session, following very minimal interaction with the research assistant. It is therefore unlikely that participants’ interactions with research assistants who knew of their assigned walk condition would have impacted observed baseline levels of negative affect. Moreover, we controlled for these baseline differences in our subsequent analyses. The presence of more female than male participants limited our ability to generalize our findings to male participants. Like other studies examining the impact of walking in nature on affect, our Pre-Walk (Time 1) measurements were assessed in a laboratory setting instead of at participants’ homes which may have impacted the accuracy of baseline measurements in reflecting true, day-to-day baseline affect. Participants in the nature walk condition had to walk through a parking lot and across a busy road for 14 minutes before reaching the nature setting. However, this may increase the generalizability of findings given that most city dwellers must travel by foot or vehicle in order to reach a natural environment.

### Clinical Implications and Conclusion

Altogether, our results suggest that walking in nature might be a useful strategy to immediately improve the affect of individuals diagnosed with MDD. The precise mechanisms by which nature exposure might influence human affect are unclear (Markevych et al., 2017), although a study based on 38 participants reported that nature exposure reduces self-reported rumination and neural activity in the subgenual prefrontal cortex, a brain structure associated with negative rumination (Bratman et al., 2015b).

Though more studies are needed to replicate findings, prior evidence suggests that wilderness therapy (e.g., Bettmann et al., 2016) and group walks in nature can serve as complementary treatment options for adults with depression (e.g., see Keenan et al., 2021 and Sturm et al., 2012). To illustrate, a crossover trial with 20 participants having attempted suicide at least once reported a reduction in depressive symptoms and hopelessness after a 9-week hiking intervention vs. a 9-week control phase (Sturm et al., 2012). More research is needed to investigate different ways in which to integrate the beneficial effects of nature exposure into existing treatment plans for adult psychiatric outpatients with major depressive disorder.

## Data Availability

The data that support the findings of this study are available on request from the corresponding author. The data are not publicly available due to privacy or ethical restrictions.

## Acknowledgements

The authors thank Alain Girard, MSc, for his contributions to the statistical analyses as well as all the participants of the study.

## Supplemental Material

**Supplemental Table 1.**
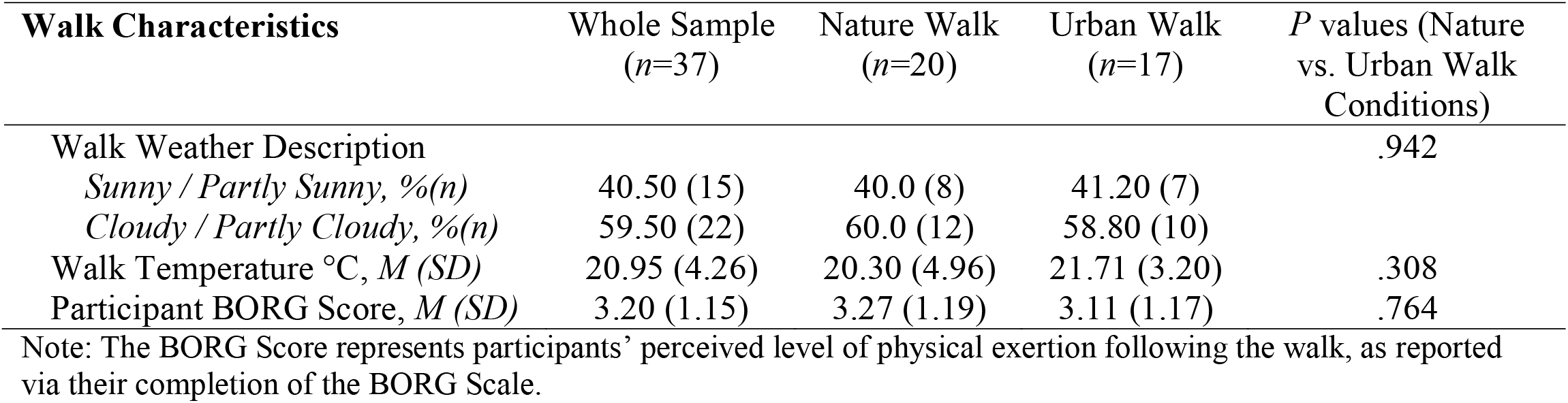
Walk characteristics in nature and urban walking conditions

### Changes in the protocol due to the COVID-19 pandemic

Data collection was temporarily halted in 2020 due to the COVID-19 pandemic. When data collection resumed in spring 2021, the following changes were made to mitigate COVID-related risks:

- The intake interview (comprising the administration of the sociodemographic questionnaire and the Hamilton Depression Rating Scale) was conducted over the phone instead of in-person;
- The collection of saliva samples for the purpose of measuring cortisol levels was removed entirely from the study’s procedure (ClinicalTrials.gov, NCT03996785).

In addition, we increased participant compensation from $50 to $75 to further incentivize in-person participation in the study in light of the fact that most patients at the DMHUI were consulting their medical teams on an entirely virtual (online) basis.

